# Absence of Excess Mortality in a Highly Vaccinated Population During the Initial Covid-19 Delta Period

**DOI:** 10.1101/2021.09.16.21263477

**Authors:** Jeremy Samuel Faust, Chengan Du, Katherine Dickerson Mayes, Benjamin Renton, Shu-Xia Li, Zhenqiu Lin, Harlan M. Krumholz

**Author notes:** corresponding author 10 Vining Street, Boston, MA 02115.

## Abstract

**Background:** All-cause excess mortality (the number of deaths that exceed projections in any period) has been widely reported during the Covid-19 pandemic. Whether excess mortality has occurred during the Delta wave is less well understood.

**Methods:** We performed an observational study using data from the Massachusetts Department of Health. Five years of US Census population data and CDC mortality statistics were applied to a seasonal autoregressive integrated moving average (sARIMA) model to project the number of expected deaths for each week of the pandemic period, including the Delta period (starting in June 2021, extending through August 28th 2021, for which mortality data are >99% complete). Weekly Covid-19 cases, Covid-19-attributed deaths, and all-cause deaths are reported. County-level excess mortality during the vaccine campaign are also reported, with weekly rates of vaccination in each county that reported 100 or more all-cause deaths during any week included in the study period.

**Results:** All-cause mortality was not observed after March 2021, by which time over 75% of persons over 65 years of age in Massachusetts had received a vaccination. Fewer deaths than expected (which we term ‘deficit mortality’) occurred both during the summer of 2020, the spring of 2021 and during the Delta wave (beginning June 13, 2021 when Delta isolates represented >10% of sequenced cases). After the initial wave in the spring of 2020, more Covid-19-attributed deaths were recorded that all-cause excess deaths, implying that Covid-19 was misattributed as the underlying cause, rather than a contributing cause of death in some cases.

**Conclusion:** In a state with high vaccination rates, excess mortality has not been recorded during the Delta period. Deficit mortality has been recorded during this period.

## Manuscript

Excess deaths—the number of all-cause fatalities exceeding the expected number in any given period—have occurred during Covid-19 outbreaks worldwide.^1^ We report that there has been no excess mortality during the Delta variant era in Massachusetts, a state with high vaccination rates, in contrast to two previous waves, and that fewer all-cause deaths than expected occurred during this period, which we term “deficit mortality.”

We applied seasonal autoregressive integrated moving average (sARIMA) models to US Census population data and CDC death statistics (2014-2019) to account for the aging population and mortality trends to project the weekly number of expected deaths in Massachusetts during the Covid-19 period (February 8, 2020-August 28, 2021), correcting for the decrease in population during the pandemic owing to cumulative excess deaths, as described elsewhere.^2,3^ The Massachusetts Department of Health supplied weekly coronavirus case counts, all-cause deaths, and Covid-19-attributed deaths (January 4 2015-August 28, 2021, >99% complete).^4^

In the first wave (March-June 2020), there were 103,150 Covid-19 cases, >7,878 Covid-19-attributed deaths (ICD-U07.1), and 7,775 excess deaths (Figure). Initially, there were more excess deaths than Covid-19-attributed deaths, perhaps owing to inadequate testing. However, recorded excess mortality in Massachusetts has been lower than Covid-19-attributed deaths since then, suggesting some misattribution (i.e. Covid-19 was likely a contributing cause, not the underlying cause of death in some cases).

**Figure.**
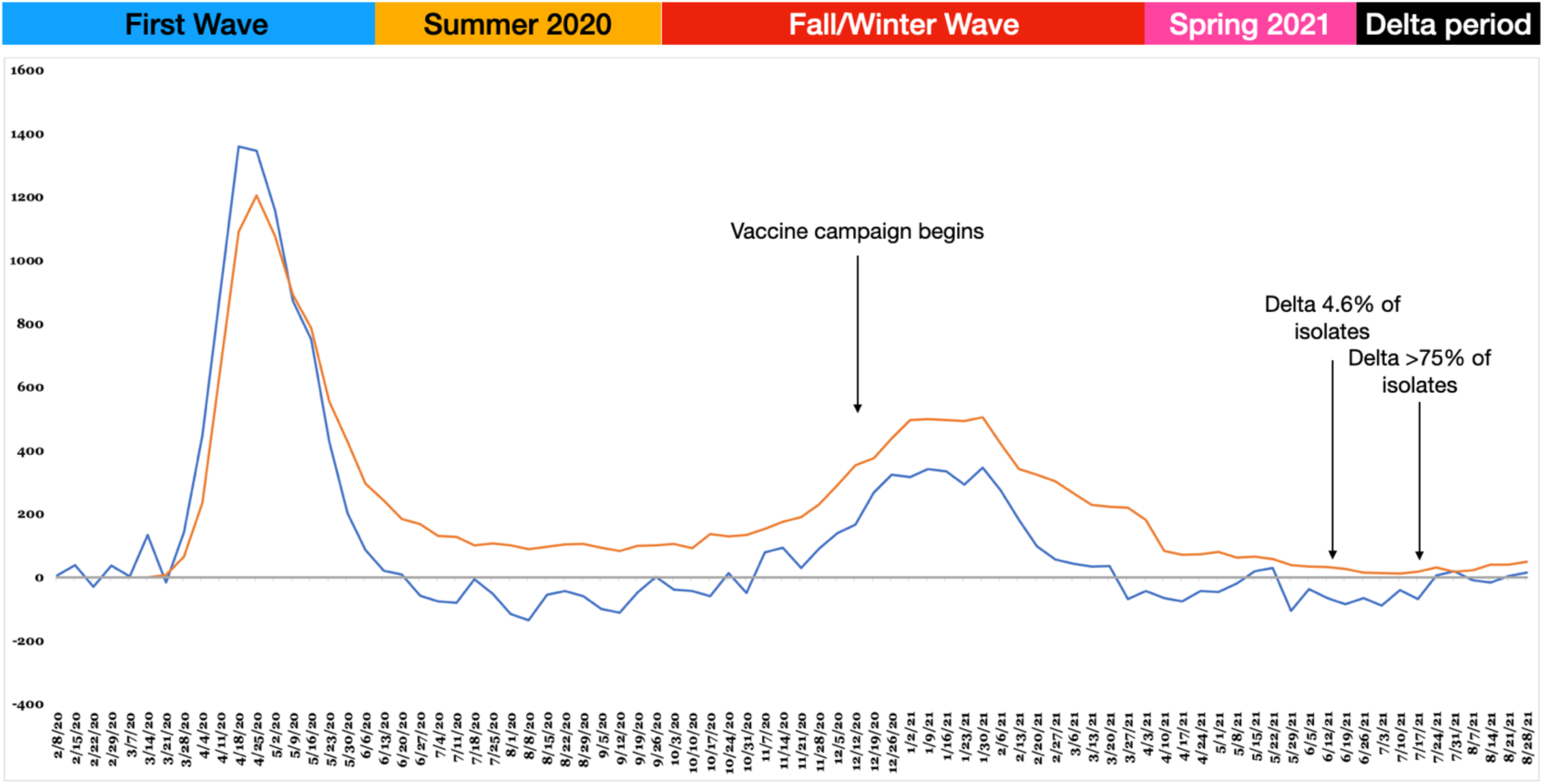
Weekly all-cause excess deaths in Massachusetts (blue line) and Covid-19-attributed deaths (orange line) in Massachusetts. The baseline (gray line) was determined using sARIMA and reflects five years of population growth, weekly mortality data, and population changes owing to the total cumulative excess deaths recorded during the pandemic.

During the summer of 2020 (July-September), there were 25,648 Covid-19 cases and >1,334 Covid-19-attributed deaths. However, 888 fewer all-cause deaths than projected occurred (“deficit mortality”) likely due to a pull-forward effect; deaths occurring months or years earlier than they would have, absent the pandemic.

In the second wave (October 2020-March 2021), there were 466,100 Covid-19 cases, 7,603 Covid-19-attributed deaths, and 3,282 excess deaths, again suggesting some underlying cause of death misattribution.

During the spring of 2021 (April 2021-June 12, 2021, the period following the second wave), deficit mortality was again observed (>460 fewer deaths than expected); 771 Covid-19-attributed deaths were reported.

During the Delta period (weeks in which Delta variants >10% of isolates in HHS Region 1, including Massachusetts,^5^ beginning the week ending June 20, 2021), there were 57,691 Covid-19 cases, and 342 Covid-19-attributed deaths; no significant excess deaths were recorded; In fact, deficit mortality persisted (337 deaths fewer occurred than expected). In the most recent two weeks (August 15-28, 2021), excess mortality >10% was observed in 2 of 8 counties (Bristol and Hampden) with large enough populations to assess (i.e. those recording ≥100 deaths during any study week), the two counties with the lowest vaccination rates among included counties (Table).

**Table.**
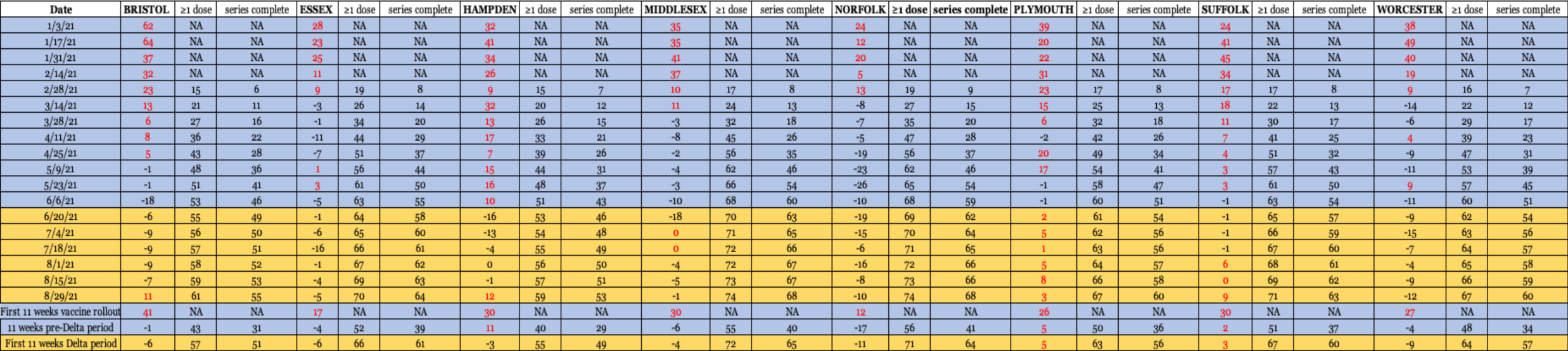
All-cause deaths (14-day periods) are shown from the start of the vaccine rollout period (the two-week period ending 1/3/2021) through the most recent week of the Delta period for which data are available. The percentage of residents in each county vaccinated for each period were obtained from the Massachusetts public vaccine dashboard.^4^

Despite increases in Covid-19 cases (including breakthrough cases), Covid-19-associated hospitalizations, and Covid-19-attriuted deaths during the Delta period, Massachusetts has not recorded state-level excess mortality since March 2021, at which time 30% of the total population and 75% of individuals ages ≥65 had received at least one dose of an authorized vaccine.^4^ This study suggests that the authorized Covid-19 vaccines remain effective in preventing severe disease and death at the population level.

## Data Availability

All data are public and available, as referenced in this work.

## References

1. Bilinski A, Emanuel EJ. COVID-19 and Excess All-Cause Mortality in the US and 18 Comparison Countries. JAMA. 2020;324(20):2100–2102. doi:10.1001/jama.2020.20717

2. Rossen LM. Disparities in Excess Mortality Associated with COVID-19 — United States, 2020. MMWR Morb Mortal Wkly Rep. 2021;70. doi:10.15585/mmwr.mm7033a2

3. Faust JS, D. C, Li S-X, Lin Z, Krumholz HM. Correcting Excess Mortality for Pandemic-Associated Population Decreases.; 2021:2021.02.10.21251461. doi:10.1101/2021.02.10.21251461

4. Massachusetts Department of Health. COVID-19 Response Reporting | Mass.gov. Accessed September 12, 2021. https://www.mass.gov/info-details/covid-19-response-reporting

5. CDC. CDC COVID Data Tracker. Accessed July 3, 2020. https://www.cdc.gov/covid-data-tracker/index.html#demographics

